# Humoral and cellular responses after a third dose of BNT162b2 vaccine in patients treated for lymphoid malignancies

**DOI:** 10.1101/2021.07.18.21260669

**Authors:** Daniel Re, Barbara Seitz-Polski, Michel Carles, Vesna Brglez, Daisy Graça, Sylvia Benzaken, Stéphane Liguori, Khaled Zahreddine, Margaux Delforge, Benjamin Verrière, Emmanuel Chamorey, Barrière Jérôme

**Author notes:** Corresponding Authors: Dr Daniel Re and Dr Jérôme Barrière **Corresponding Author:** Dr Barrière Jérôme, Department of medical oncology, Clinique Saint Jean, 92 avenue Dr Donat, 06800 CAGNES-SUR-MER, FRANCE. Authors contributed equally to this work. **Author Contributions:** Drs Re, Seitz-Polski, Chamorey and Barrière had full access to all of the data in the study and take responsibility for the integrity of the data and the accuracy of the data analysis. Concept and design: All authors. Acquisition, analysis, or interpretation of data: All authors. Drafting of the manuscript: Drs Re, Seitz-Polski, Carles, Chamorey and Barrière. Critical revision of the manuscript for important intellectual content: All authors. Statistical analysis: Dr Seitz-Polski and Chamorey. Administrative, technical, or material support: All authors. Supervision: Drs Re and Dr Barriere. **Funding/Support:** This research was supported by a grant from Conseil Départemental des Alpes-Maritimes and from the Agence Nationale pour la Recherche AO-Flash-COVID. **Role of the Funder/Sponsor:** The funders had no role in the design and conduct of the study; collection, management, analysis, and interpretation of the data; preparation, review, or approval of the manuscript; and decision to submit the manuscript for publication. **Disclaimer:** The analyses described are the responsibility of the authors and do not necessarily reflect the views or policies of the US Department of Health and Human Services. The mention of trade names, commercial products, or organizations does not imply endorsement by the US government. **Additional Contributions:** We acknowledge all nurses of the day clinic of the Antibes Hospital (Elodie, Laurence, Nicole, Vinaj, and Valérie) and more specifically Sylvie Andreo who was responable for the administration of dose 3 and for the management of blood sampling. We last acknowledge patients for their participation.

## Abstract

**BACKGROUND:** Immunocompromised patients such as patients with hematological malignancies have impaired immune response to two doses of BNT162b2 (Pfizer / BioNtech) vaccine against SARS-CoV-2. Evaluation of a repeated immune stimulation with a third vaccine dose is needed.

**METHODS:** a vaccine monitoring observatory was conducted in outpatients who were treated for lymphoid malignancies (LM) to monitor both immune and cellular response measured the day of administration of the dose 3 of the mRNA vaccine BNT162b2 and again three to four weeks. Elecsys ® Anti-SARS-CoV-2 immunoassay was used to asses to the level of SARS-CoV-2 anti-Spike (S) antibodies (Abs) titer and SARS-CoV-2-specific T-cell responses were assessed by a whole blood Interferon-Gamma Release Immuno Assay (IGRA) (QuantiFERON Human IFN-gamma SARS-CoV-2, Qiagen®).

**RESULTS:** Among the 43 assessable patients (suffering from chronic lymphocytic leukemia (CLL) (n=15), indolent and aggressive B cell non-Hodgkin lymphoma (NHL) (n=14), and multiple myeloma (MM) (n=16)), 18 (41,8%) had no anti-S Abs before the dose 3 of BNT162b2 vaccine (n=9 CLL, n=8 NHL, n=1 MM), and they all 18 remained negative after the dose 3. Amongst the 25 patients with positive anti-S titers before dose 3, all patients remained positive and 23 patients increased their anti-S titer after dose 3. Patients with CLL and/or with previous anti-CD20 therapy treated within 12 months of administration of dose 3 had no significant increase of the humoral response. Among 22 available patients, dose 3 of BNT162b2 vaccine significantly increased the median IFN-gamma secretion. On eight (36.4%) patients who were double-negative for both immune and cellular response, five (22.7%) patients remained double-negative after dose 3.

**CONCLUSIONS:** Dose 3 of BNT162b2 vaccine stimulated humoral immune response among patients with LM, in particular patients with MM (who had higher anti-S baseline titer after dose 2) and those with no anti-CD20 treatment history within a year. T-cell response was increased among patients in particular with no active chemotherapy regimen. Our data support the use of an early third vaccine dose among immunocompromised patients followed for LM though some of them will still have vaccine failure.

## Manuscript

Patients suffering from solid cancer (SC) or hematological malignancies (HM) are at increased risk of severe Coronavirus disease (COVID-19) (1-2), caused by infection with severe acute respiratory syndrome coronavirus 2 (SARS-CoV-2). Vaccination against SARS-CoV-2 induces high rates of seroconversion in a healthy population but is less effective in immunocompromised patients such as organ transplanted patients (3), patients treated for SC (4-6) or patients with HM (7-9). We and others showed previously that patients treated with anti-CD20 monoclonal antibodies (Mabs) prior to COVID-19 vaccination had a very low likelihood of developing a humoral response, especially if the anti-CD20 Mab treatment is performed within 6 to 12 months of administration of the anti-SARS-CoV-2 vaccine (8-9). T-cell response seemed more impaired in patients with HM than with SC, with a positive effect of the booster dose (6).

We conducted a specific analysis of patients followed at our hospital for lymphoid malignancies (LM), who showed no or poor humoral response after vaccination with two doses of the mRNA vaccine BNT162b2 (Pfizer / BioNtech). We monitored their humoral and cellular response, after a third dose (dose 3) of BNT162b2 according to the recommendation of the French National Authority for health (10) published on April 11^th^ 2021.

## Methods

All participants signed a written informed consent and accepted their participation in this registered trial in accordance with ethical and legal French policies (Registration number F20210324145532). All data were prospectively collected on an electronic anonymous data set. Humoral responses were measured with Elecsys ® Anti-SARS-CoV-2 immunoassay (Roche Diagnostics, France) with detection of antibodies (Abs) directed to total Abs (immunoglobulin (Ig) G, IgA, IgM) against the SARS-CoV-2 Nucleocapsid (N) antigen (qualitative detection) and total Abs against the SARS-CoV-2 Spike (S) protein receptor binding domain (quantitative detection). For the anti-N assay, serum showing an index ≥ 1.0 was considered to be reactive and suggested a potential virus contact. For the anti-S assay, serum showing a result ≥ 0.8 U/mL was declared positive and suggested an effective immune response virus- or vaccine-related. These assays were a double-antigen sandwich electrochemiluminescence and were performed on an a Cobas e 601 automate (Roche). The level of SARS-CoV-2 anti-N and anti-S Abs was measured the day of administration of the dose 3 of the mRNA vaccine BNT162b2 and again three to four weeks later (median 27 days (d), range [21; 35]).

SARS-CoV-2-specific T-cell responses were assessed by a whole blood Interferon-Gamma (IFN-gamma) Release Immuno Assay (IGRA) using two Qiagen® proprietary mixes of SARS-CoV-2 S protein (antigen 1 and antigen 2) selected to activate both CD4 and CD8 T cells. Briefly, venous blood samples were collected directly into the Quantiferon® tubes containing either spike peptides or positive and negative controls. Whole blood was incubated at 37°C for 16-24 hours and centrifuged to separate plasma. Interferon gamma was measured in these plasma samples using enzyme-linked immunosorbent assay tests (QuantiFERON Human IFN-gamma SARS-CoV-2, Qiagen®) (11).

Data are presented as median and range for continuous values with non-Gaussian distribution. The D’Agostino & Pearson normality test was used to determine if a variable had a Gaussian distribution or not. Continuous values were normalized using logarithm function when appropriate. Continuous values were compared by Student T test or Mann-Whitney test when appropriate. Categorical data were summarized using counts and percentages; they were compared by using Chi2 test or Fisher exact test when appropriate. Statistical analyses were performed using R.4.0.3 and GraphPad Prism 7.0 (GraphPad Software, Inc., San Diego, CA). All comparisons were two-tailed, and the differences were considered significant when P-value < 0.05.

## Results

We analyzed a data set of 45 patients, prospectively included to receive dose 3 of the BNT162b2 vaccine given 78 days [range: 47-114] after dose 2 of the same vaccine. All 45 patients were negative for anti-N Abs before dose 3, but two patients were tested positive after dose 3 and were therefore excluded from the final analysis. Included patients were suffering from chronic lymphocytic leukemia (CLL) (n=15), indolent and aggressive B cell non-Hodgkin lymphoma (NHL) (n=14), and multiple myeloma (MM) (n=16). The median age of the 43 patients included in the final analysis was 77 years [range: 37-92], 63 % were men and 37 % women (Table 1). At the time of dose 3, patients’ treatments are reported in Table 1.

**Table 1:**
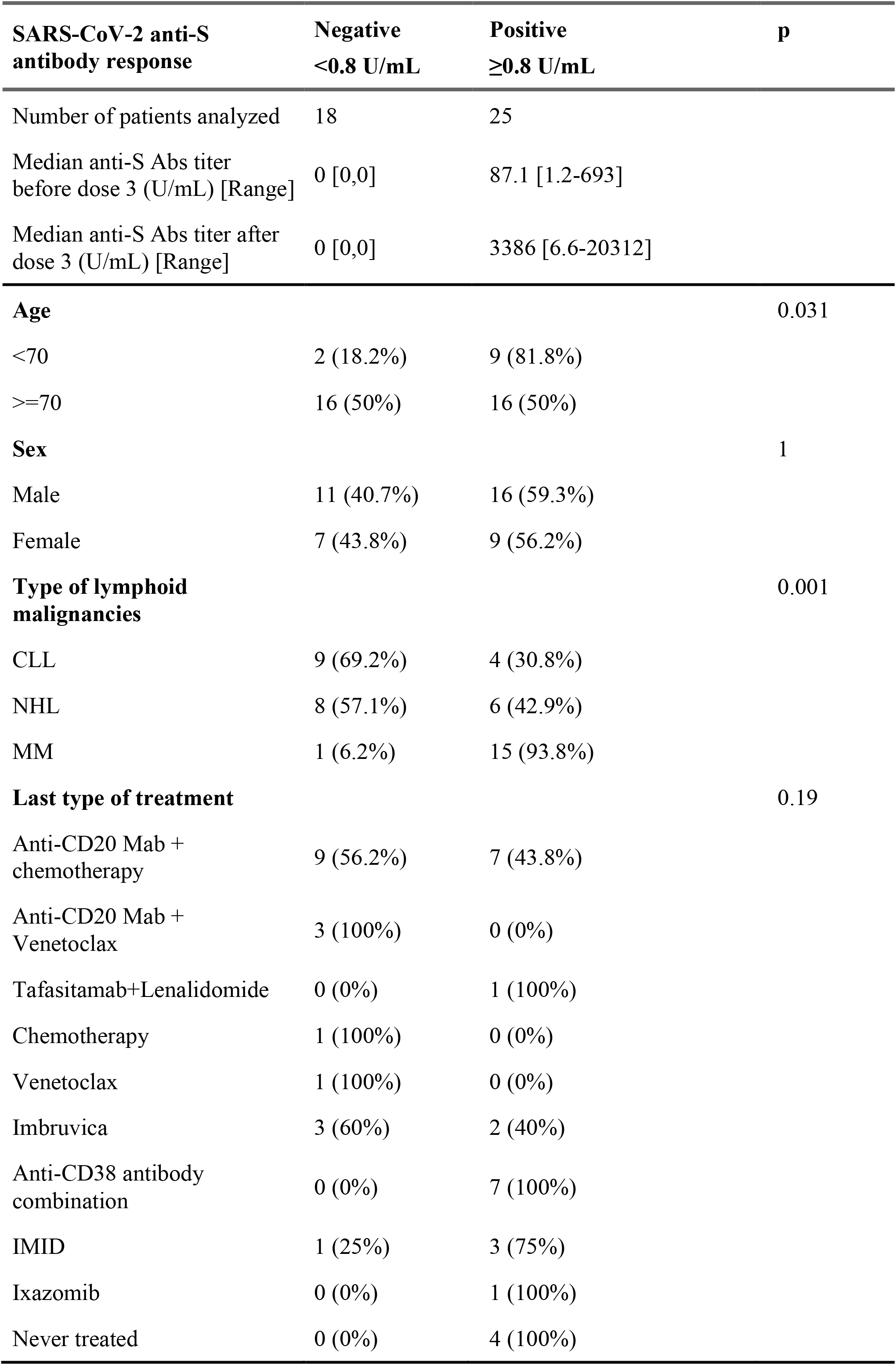
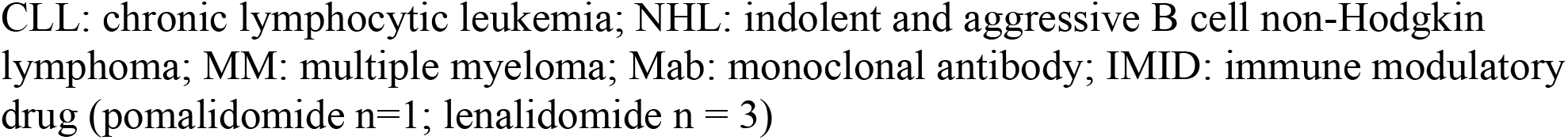
Characteristics of patients included in this study, before and after dose 3 (same data).

| **SARS-CoV-2 anti-S antibody response** | **Negative**<br>**<0.8 U/mL** | **Positive**<br>**≥0.8 U/mL** | **p** |
| --- | --- | --- | --- |
| Number of patients analyzed | 18 | 25 |  |
| Median anti-S Abs titer before dose 3 (U/mL) [Range] | 0 [0,0] | 87.1 [1.2-693] |  |
| Median anti-S Abs titer after dose 3 (U/mL) [Range] | 0 [0,0] | 3386 [6.6-20312] |  |
| **Age** |  |  | 0.031 |
| <70 | 2 (18.2%) | 9 (81.8%) |  |
| ≥70 | 16 (50%) | 16 (50%) |  |
| **Sex** |  |  | 1 |
| Male | 11 (40.7%) | 16 (59.3%) |  |
| Female | 7 (43.8%) | 9 (56.2%) |  |
| **Type of lymphoid malignancies** |  |  | 0.001 |
| CLL | 9 (69.2%) | 4 (30.8%) |  |
| NHL | 8 (57.1%) | 6 (42.9%) |  |
| MM | 1 (6.2%) | 15 (93.8%) |  |
| **Last type of treatment** |  |  | 0.19 |
| Anti-CD20 Mab + chemotherapy | 9 (56.2%) | 7 (43.8%) |  |
| Anti-CD20 Mab + Venetoclax | 3 (100%) | 0 (0%) |  |
| Tafasitamab+Lenalidomide | 0 (0%) | 1 (100%) |  |
| Chemotherapy | 1 (100%) | 0 (0%) |  |
| Venetoclax | 1 (100%) | 0 (0%) |  |
| Imbruvica | 3 (60%) | 2 (40%) |  |
| Anti-CD38 antibody combination | 0 (0%) | 7 (100%) |  |
| IMiD | 1 (25%) | 3 (75%) |  |
| Ixazomib | 0 (0%) | 1 (100%) |  |
| Never treated | 0 (0%) | 4 (100%) |  |

CLL: chronic lymphocytic leukemia; NHL: indolent and aggressive B cell non-Hodgkin lymphoma; MM: multiple myeloma; Mab: monoclonal antibody; IMiD: immune modulatory drug (pomalidomide n=1; lenalidomide n = 3)

### Humoral immunity

Amongst 43 patients, 18 (41,8%) had no anti-S Abs before the dose 3 of BNT162b2 vaccine (n=9 CLL, n=8 NHL, n=1 MM), and they all 18 remained negative after the dose 3 (Table 1). Fourteen of these 18 patients had already received an anti-CD20 Mab treatment, nine of them within the 12 months preceding the vaccine. One seronegative patient with MM was under active treatment for HIV infection.

In univariate analysis, age and type of LM, but not sex or type of treatment (except for anti-CD20 Mab performed within 6 to 12 months of administration of the anti-SARS-CoV-2 vaccine), was statistically associated with anti-S response after dose 3 (Table 1).

Amongst the 25 patients with positive anti-S titers before dose 3, all patients remained positive (100%) and 23 patients increased their anti-S titer after dose 3. Their median anti-S titer increased from 87.1 U/mL [range: 1.2-693] to 3386 U/mL [range: 6.6-20312] (p < 0.001) (Table 1). The median anti-S titer changed as following: 0 U/mL [range: 0-120] to 0 U/mL [range: 0-5997] (p = 0.12) in patients with CLL, from 0 [range: 0-310] to 0 U/mL [range: 0-6101] (p = 0.07) in patients with NHL, and from 100 U/mL [range: 0-690] to 2700 U/mL [range: 0-20312] (p < 0.0001) in patients with MM (Figure 1A). We tested the impact of previous anti-CD20 Mab therapy and found that patients treated within 12 months of administration of dose 3 did responded poorly (median anti-S titer: 0 U/mL [range: 0-6101]) when compared to patients that received the same drug at least 12 months before BNT162b2 vaccine (median anti-S titer: 4200 [range: 0-6073]) (p=0.047) (Figure 1B).

**Figure 1A:**
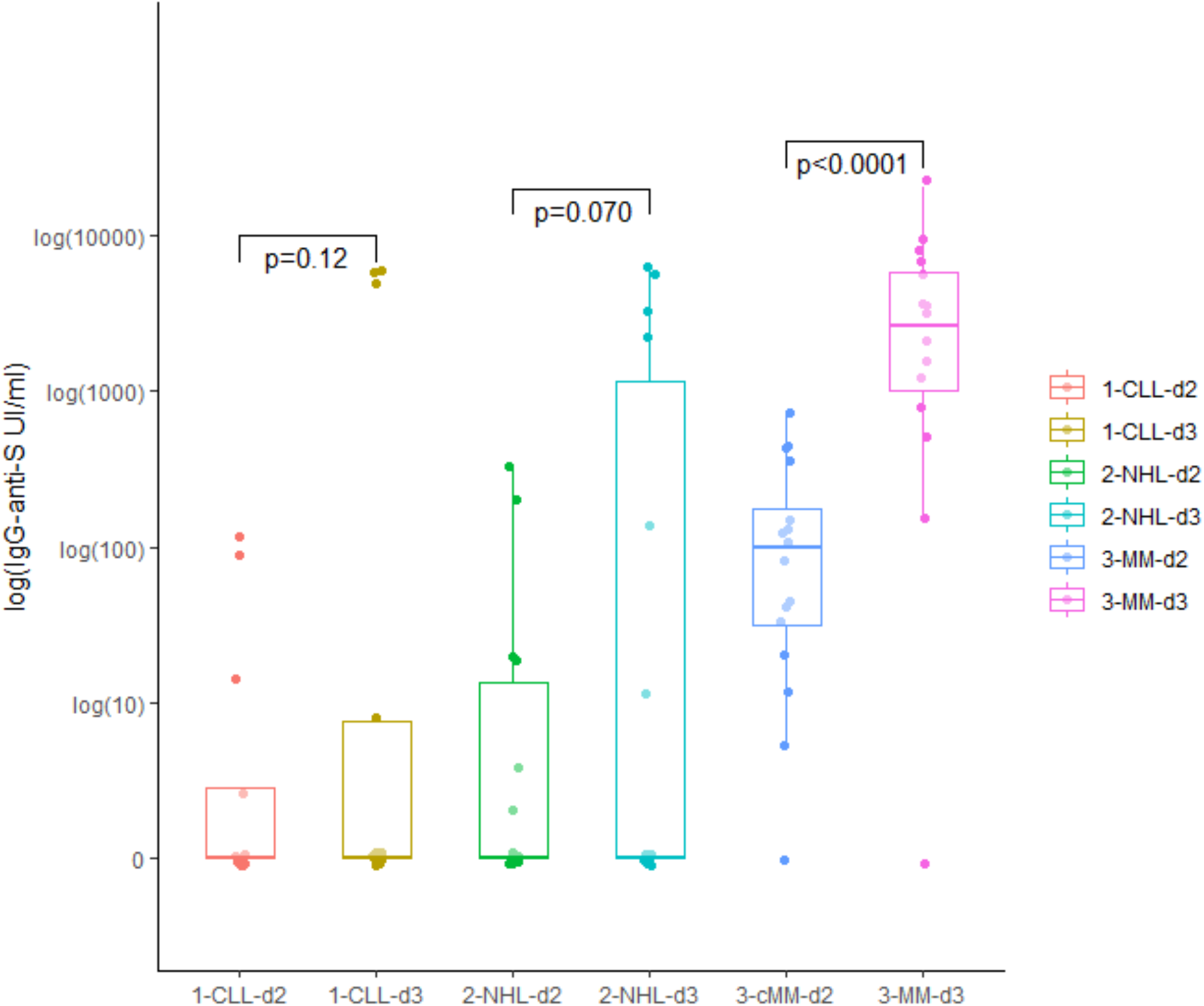
Humoral quantitative anti-Spike (S) antibodies (logarithmic scale) response after dose 2 (d2) and after dose 3 (d3) of the BNT162b2 vaccine in 43 patients with lymphoid malignancies (n = 13 patients with chronic lymphocytic leukemia (CLL), n = 14 patients with B cell non-Hodgkin lymphoma (NHL) and n = 16 patients with multiple myeloma (MM))

**Figure 1B.**
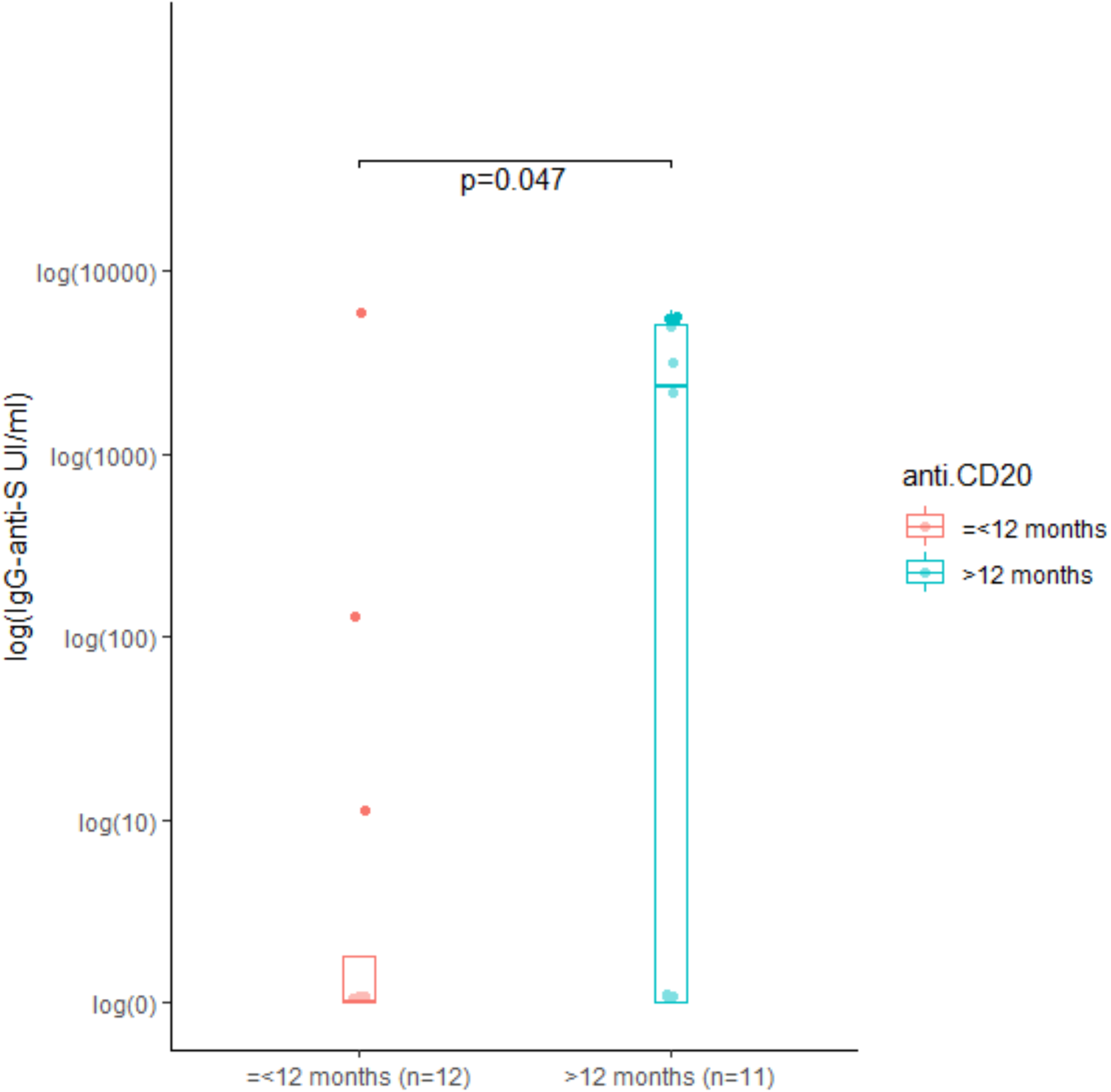
Humoral quantitative anti-Spike (S) antibodies (logarithmic scale) after dose 3 of the BNT162b2 vaccine in 23 patients pre-treated with an anti-CD20 Mab within <= 12 months (n = 12) or prior to > 12 months (n = 11).

### Exploratory comparative cellular immunity response with humoral response

We next aimed to characterize specific T-cell responses to the BNT162b2 vaccine with a whole blood IGRA test in 27 patients with CD-20 positive disease to assess comparative immune response among poor humoral immune responders. We were able to collect and analyze blood samples in 22 (CLL n=10, NHL n=12) of them.

After dose 2, 15 patients showed a positivity for both humoral and T-cell response, seven patients were positive for only one of the two tests. Of note, four patients showed a T-cell response despite absence of measurable anti-S Abs. A total of eight patients (36.4%) were double-negative for both tests.

Dose 3 of BNT162b2 vaccine increased median IFN-gamma secretion after exposition to antigen 1 or antigen 2 (from 0.07 IU/ml [range: 0.0-0.17] to 0.3 IU/mL [range: 0.0-0.9] (p=0.0008) and from 0.06 IU/ml [range: 0.0-0.1] to 0.2 IU/mL [range: 0.0-1.3] (p=0.0006), respectively) (Figure 2A). Three patients that were initially double-negative showed a T-cell response, leaving a total of five patients (22.7%) without B- or T-cell response after dose 3. These patients included three cases of CLL (all actively treated with venetoclax and a past history of rituximab treatment) and two cases of NHL (rituximab and bendamustine, one on and one off this treatment combination). Four of those five double-negative patients had thus active treatment.

**Figure 2.**
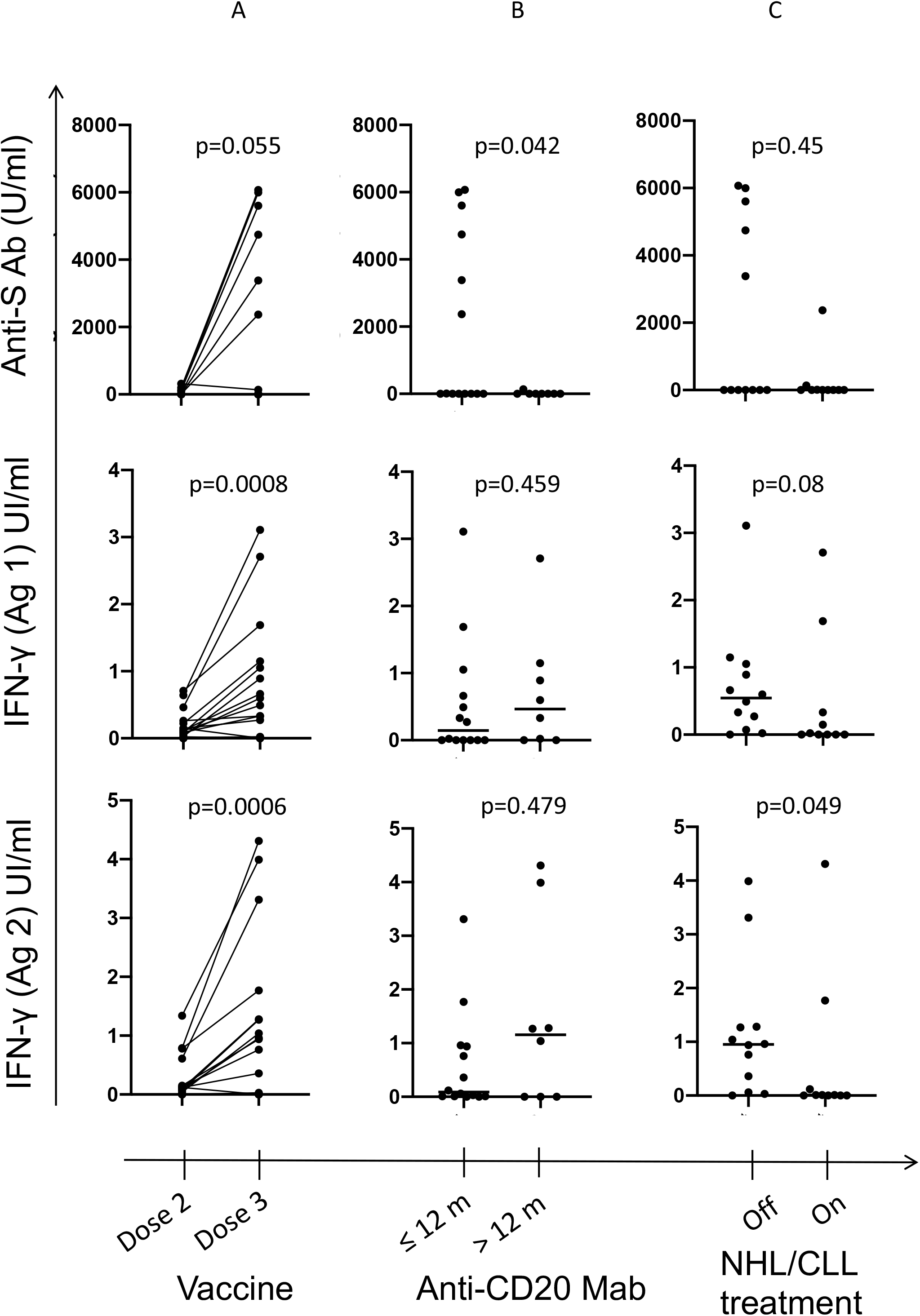
**A** : Anti-S Ab titer and IFN-gamma secretion after exposition to antigen 1 or antigen 2 in 22 patients with CLL and NHL before and 25 days after administration of the BNT162b2 vaccine (dose 2 and 3, respectively). Dose 3 increased IFN-gamma secretion after exposition to antigen 1 or antigen 2 (p=0.0008 and p=0.0006, respectively) **B** : Anti-S Ab titer and IFN-gamma secretion after exposition to antigen 1 or antigen 2 in 22 patients with CLL and NHL depending on the delay between of last administration of anti-CD20 treatment and administration of the BNT162b2 vaccine. The median (range) of IFN-gamma secretion after exposition to antigen 1 or antigen 2 was 0.14 IU/mL [0.0 −3.1] and 0.09 IU/mL [0.0 - 0.9] for patients pre-treated prior to > 12 months vs 0.5 IU/mL [0.0-1.1] and 1.2 IU/mL (0.0-3.3] for patients pre-treated within <= 12 months (p=0.45 and p=0.48 respectively). **C** : Anti-S Ab titer and IFN-gamma secretion after exposition to antigen 1 or antigen 2 in 22 patients with CLL and NHL depending on the administration of an ongoing active treatment during the vaccination sequence with BNT162b2. The median (range) of IFN-gamma secretion after exposition to antigen 1 was 0.0 IU/mL [0.0-0.7] for patients with active treatment vs. 0.5 IU/mL [0.1; 1.0] for patients without active treatment (p=0.08) and to antigen 2 was respectively 0.0 IU/mL [0.0-0.5] vs 0.9 IU/mL [0.1; 4.0] (p=0.049).

While the timing of treatment with an anti-CD20 Mab impacted humoral response as already stated, it did not modify T-cell response: median of IFN-gamma secretion after exposition to antigen 1 or antigen 2 was 0.14 IU/ml [range: 0.0-3.1] and 0.09 IU/mL [range: 0.0-0.9], respectively, for patients treated prior to > 12 months versus 0.5 IU/mL [range: 0.0-1.1] and 1.2 IU/mL [range: 0.0-3.3], respectively, for patients treated within <= 12 months (p=0.45 and p=0.48 respectively) (Figure 2B). Patients on active NHL or CLL treatment during the vaccination sequence had a poorer specific T-cell response than patients without ongoing cancer specific medication, the median of IFN-gamma secretion after exposition to antigen 2 was 0.0 IU/ml [range: 0.0; 0.5] vs 0.9 IU/mL [range: 0.1-4.0] p=0.049 (p = 0.08 for antigen 1) (Figure 2C). There was no difference of T-cell response between patients with CLL or NHL (p>0.99).

We then compared the performance of serologic and IGRA testing using the method of the receiver operating characteristic (ROC) curve to explore the best way to detect a specific immune response to SARS-CoV-2 vaccine. In this cohort of 22 patients (and seven healthy subjects naïve for SARS-Cov2 infection as negative controls), the IGRA test based on two different antigens identified more efficiently than the Elecsys ® Anti-SARS-CoV-2 immunoassay a specific immune response to SARS-CoV-2 vaccine (aera under the curve (AUC) anti-S Abs: 0.7045, p=0.11; AUC IGRA Antigen 1: 0.8636, p = 0.004 (Sensitivity: 65%, Specificity: 100%); AUC IGRA Antigen 2 : 0.8864, p=0.002, (Sensitivity: 60%, Specificity: 100%)).

### Tolerance of the dose 3

No novel adverse events were observed in our population after dose 3 of BNT162b2 vaccine.

## Discussion

To our knowledge, this study is the first to report results on cellular and humoral immunity after administration of a third dose of BNT162b2 vaccine in patients treated for LM. To date, only one publication has demonstrated a favorable impact of the administration of a third dose of BNT162b2 vaccine to solid-organ transplant recipients with a significantly improved humoral response (12).

Previous reports highlighted reduced rates of seroconversion after two doses of a SARS-CoV-2 vaccine for patients with HM, leaving some of them without detectable anti-S Ab protection (7-9), with lower response for patients with CLL, even without treatment, or patients under anti-CD20 therapy. We here showed that patients without Ab response after two doses do not benefit from a third dose of the same vaccine when the analysis is limited to serologic testing, thus eliminating the need of anti-S level measurement before and after a third vaccination in current clinical practice. The third vaccine dose increased the overall humoral response in those patients responding after the second dose, especially patients with MM, and to a lower extent patient with NHL, but not patients with CLL, suggesting a negative impact of the pathology on the induction of a specific T-cell response.

Similar to patients with anti-CD20 treated multiple sclerosis (13), some of our patients with LM did not develop a specific T cell response to the second dose. In contrast, several of the seronegative patients showed an emerging cellular response after dose 3, suggesting a positive effect of a second booster dose despite the absence of humoral response. Given the importance of a T-cell response in critically ill COVID-19 patients (14-15), this is another expected benefit of the dose 3 vaccine dose, especially for patients treated with anti-CD20 Mab based therapy who are at high risk for death or ongoing SARS-CoV-2 shedding (16-17).

Our study has some limitations, firstly due to the small number of included patients. Then, we were unable to measure the level of neutralizing Abs (availability and cost issues), preventing any correlation assessment of these Abs and the level of anti-S Abs. Finally, only a subgroup of the cohort has been evaluated for the cellular immunity, due to availability issues: a larger sample would have allowed more detailed subgroup analyzes, in particular according to the different treatments received.

Considering the decrease of humoral immunity over time (18), the known correlation between neutralizing Abs titers and clinical response (19), and the here shown emergence of a specific cellular T-cell response, an early third dose of the SARS-CoV-2 vaccine seems necessary in patients with LM to allow optimal protection, even if anti-CD20 treated patients and patients treated with stem cell toxic drugs such as bendamustine remains a major concern. For these patients with vaccine failure, in addition to the incentive of relatives to get vaccinated and the drastic maintenance of social protection measures, repeated immune stimulation with a fourth vaccine dose, a multimodal immune stimulation with heterologous prime-boost vaccination, or a maximized immune stimulation double-dose approach should be considered (20).

## Data Availability

all data are available

## Notes

**Conflict of Interest Disclosures:** No disclosures were reported.

### Competing Interest Statement

The authors have declared no competing interest.

### Clinical Trial

All participants signed a written informed consent and accepted their participation in this registered trial in accordance with ethical and legal French policies (Registration number F20210324145532)

### Funding Statement

This research was supported by a grant from Conseil Departemental des Alpes-Maritimes and from the Agence Nationale pour la Recherche AO-Flash-COVID.

### Author Declarations

The third dose was made according to the French National Authority for health (ref 10) published on April 11th 2021 recommending the use of a third dose in immunosuppressed patients. The current study has been discussed with the president of the local ethics committee of the Antibes hospital where all patients were followed and vaccinated (Dr Philippe de Swardt). Dr de Swardt decided that the current observatory was conducted according to the French policies and therefore that a formal local or national CPP (Comite de protection des personnes) statement was not necessary according to French policies. However, all participants signed a written informed consent and accepted their participation in this registered observatory in accordance with ethical and legal French policies (Registration number F20210324145532). All data were prospectively collected on an electronic anonymous data set.

